# Impact of Vaccination Rates, Pre-Pandemic Life Expectancy, Economic Status and Age on COVID-19 Excess Mortality Across United States

**DOI:** 10.1101/2024.01.21.24301582

**Authors:** Olga Matveeva, Aleksey Y. Ogurtsov, Svetlana A. Shabalina

## Abstract

**Aim:** This study investigates factors influencing pandemic mortality rates across U.S. states during different waves of SARS-CoV-2 infection from February 2020 to April 2023, given that over one million people died from COVID-19 in the country.

**Methods:** We performed statistical analyses and used linear regression models to estimate age-adjusted and unadjusted excess mortality as functions of life expectancy, vaccination rates, and GDP per capita in U.S. states.

**Results and Discussion:** States with lower life expectancy and lower GDP per capita experienced significantly higher mortality rates during the pandemic, underscoring the critical role of underlying health conditions and healthcare infrastructure, as reflected in these factors. When categorizing states by vaccination rates, significant differences in GDP per capita and pre-pandemic life expectancy emerged between states with lower and higher vaccination rates, likely explaining mortality disparities before mass vaccination. During the Delta and Omicron BA.1 waves, when vaccines were widely available, the mortality gap widened, and states with lower vaccination rates experienced nearly double the mortality compared to states with higher vaccination rates (Odds Ratio 1.8, 95% CI 1.7-1.9, p < 0.01). This disparity disappeared during the later Omicron variants, likely because the levels of combined immunity from vaccination and widespread infection across state populations became comparable. We showed that vaccination rates were the only significant factor influencing age-adjusted mortality, highlighting the substantial impact of age-specific demographics on both life expectancy and GDP across states.

**Conclusion:** The study underscores the critical role of high vaccination rates in reducing excess deaths across all states, regardless of economic status. Vaccination rates proved more decisive than GDP per capita in reducing excess deaths. Additionally, states with lower pre-pandemic life expectancy faced greater challenges, reflecting the combined effects of healthcare quality, demographic variations, and social determinants of health. These findings call for comprehensive public health strategies that address both immediate interventions, like vaccination, and long-term improvements in healthcare infrastructure and social conditions.

## Introduction

In May 2023, the U.S. Centers for Disease Control and Prevention (CDC) and the World Health Organization (WHO) officially declared the end of the COVID-19 pandemic as a public health emergency. However, despite the decline in COVID-19 infections, persistent morbidity, and mortality, coupled with the looming threat of future significant outbreaks, necessitate a thorough examination of the effectiveness of past pandemic control strategies. Numerous studies confirm the effectiveness of COVID-19 vaccination in reducing deaths across US states (1–4). State-level analyses reveal that high vaccination rates are associated with a lower risk of hospitalization due to COVID-19 (4) and fewer overall hospitalizations (2) estimated that vaccination prevented roughly 5,000 hospitalizations and saved approximately 700 lives per 100,000 people in the US (5).

A sustained reduction in mortality rates, along with an increase in vaccination coverage, has been observed across states throughout the Alpha, Delta, and Omicron waves of the pandemic (6). Notably, a 10% increase in vaccination coverage was linked to an 8% decrease in mortality and a 7% decrease in COVID-19 cases (7). It has been observed that the 10 states with the highest vaccination rates had all-cause mortality rates comparable to, or even lower than, some European countries during the combined Delta and Omicron waves (3). Thus, research suggests significant geographical disparities in vaccination coverage across the US, with areas of low vaccination identified as “cold spots” (8). This unequal access to vaccination may be linked to variations in healthcare capacity.

The research team led by Bollyky et al. (9) conducted a comprehensive analysis of the factors influencing the impact of the COVID-19 pandemic in the US. They found that socioeconomic factors such as low poverty levels, higher education, and trust in institutions were linked to lower death rates. Conversely, states with higher percentages of minority populations experienced higher death rates. Access to quality healthcare was shown to reduce deaths and infections, while increased public health spending and personnel did not significantly affect outcomes.

While the Bollyky et al. (2023) study (9) found no direct correlation between state Gross Domestic Product (GDP) per capita and pandemic age-adjusted mortality within the US, lower GDP is often associated with challenges in healthcare infrastructure, resource availability, and social support systems. These limitations may indirectly contribute to higher overall pandemic mortality rates. Globally, studies have shown a link between low excess mortality, high vaccination rates, and high GDP (10–12). However, the complex interplay of these factors and the mechanisms by which they affected pandemic mortality in the US context remains understudied. Our comprehensive analysis across all 50 states addresses this gap by examining the relationship between age-adjusted and unadjusted pandemic mortality estimates, vaccination rates, and socioeconomic factors (including GDP per capita and pre-pandemic life expectancy).

For this analysis, we chose to assess excess mortality because it provides a more comprehensive measure of the overall impact of the pandemic than reported COVID-19 deaths alone. It accounts for both direct COVID-19 fatalities and indirect effects such as disruptions in healthcare systems, delayed medical treatments, and other socioeconomic impacts.

We also analyzed the relationship between pre-pandemic life expectancy and pandemic mortality. So far, we have been unable to find published studies that directly investigate this relationship and the mechanisms underlying it. The main research focus of published research is on addressing the question of how the pandemic has shortened life expectancy in different jurisdictions internationally (13) and within the USA (14). However, accounting for state-specific pre-pandemic life expectancy when analyzing pandemic mortality is important because life expectancy captures and integrates the impact of underlying health conditions, healthcare quality, and demographic differences, such as age distribution, which influence vulnerability to pandemics. It also highlights differences in healthcare infrastructure and social determinants of health, such as income and access to care, which can exacerbate mortality rates.

This study takes statistical approaches by simultaneously considering many factors in univariable and multivariable regression analysis along with partial correlation estimates.

By integrating both vaccination rates and economic indicators, this research seeks to shed light on the intricate dynamics influencing excess mortality, paving the way for more effective and targeted public health interventions. Furthermore, investigating the relationship between pandemic mortality and life expectancy across U.S. states can yield valuable insights into how the pandemic has disproportionately impacted different populations within the United States.

## Materials and Methods

### Life expectancy

Life expectancy at birth estimated for 2019 for US states was extracted from US Mortality Database (15).

### Total pandemic age-adjusted and unadjusted pandemic mortality from the earlier published study

Age-adjusted and unadjusted COVID-19 pandemic mortality estimates for each U.S. state, spanning January 1, 2020, to July 31, 2022, were derived from a previously published study (9). In addition we also used excess mortality estimates (age-unadjusted) derived form CDC modelling as desribed below.

### Excess mortality derived from CDC modeling

Data on excess mortality in the United States were also sourced from the US Centers for Disease Control and Prevention (CDC) (16). We downloaded the file “National and State Estimates of Excess Deaths” for our analysis. To ensure proper comparisons, we normalized the absolute values of excess mortality for each specific period using the population size of each state obtained from the United States Census Bureau’s 2020 data (17). For this normalization process, we used the file “Annual Estimates of the Resident Population for the United States, Regions, States, District of Columbia, and Puerto Rico: April 1, 2020, to July 1, 2022.”

CDC’s method for calculating excess deaths involves a modeling approach that compares recent mortality data to historical trends dating back to 2013 (18). This analysis aims to identify significant increases in all-cause mortality.

### Vaccination dataset

Vaccination data for the United States was also procured from the US Centers for Disease Control and Prevention (CDC) (19). We downloaded the file “COVID-19 Vaccinations in the United States, Jurisdiction” and extracted data that were relevant to the dates October 2, 2021, and January 2, 2022, from the column “titled “Series_Complete_Pop_Pct” and booster doses from the column titled “Additional_Doses_Vax_Pct”. Information about the file is available in CDC (19). Additionally, we included a dataset on the state’s annual per capita gross domestic product (GDP) per capita for 2020 and 2021. We obtained this dataset from the Bureau of Economic Analysis (20), and used the file “Annual GDP by State and Industry”.

### Vaccination hesitancy

Data were obtained from Office of Assistant Secretary for Planning and Evaluation (ASPE) (21). The file titled “Predicted-Vaccine Hesitancy-by State, PUMA, and County” was used for the study.

### Statistical analysis

The time intervals for analysis were classified as follows:

I. Pre-Delta (February 1, 2020-June 30, 2021): This period represents the initial period of the pandemic before the emergence of the Delta variant.
II. Delta (July 1, 2021-January 31, 2022): The Delta variant was predominant, and mass vaccination was already underway. The Delta-Omicron transition happened in December/January 2022.
III. Omicron BA.1 (February 1, 2022-March 30, 2022): The specific period corresponds to prevalence of the first Omicron variant.
IV. Post-Omicron BA.1 (April 1, 2022-April 1, 2023): This interval covered the period after the emergence of the first Omicron variant. Summary information for statistical analysis from the specified databases for the indicated time intervals was presented in Supplementary Table 1.

All states were categorized based on real GDP per-capita in 2020 using the threshold that was chosen arbitrarily. States with GDP per-capita above $65,000 were classified as higher-income, while those below $65,000 were considered less affluent.

Statistical analysis, including Pearson correlation analysis, linear regression fitting, and Odds Ratio calculations, was performed using Excel functions. In addition, we validated the results using the online tool (22).

## Results

### Complex interplay between pandemic mortality, life expectancy, GDP, and vaccination rates across US States

To understand the impact of age on excess pandemic mortality across U.S. states, we analyzed the relationship between age-adjusted and unadjusted estimates. We correlated these estimates with each other, with pre-pandemic state-specific life expectancy and annual GDP per capita values. Interestingly, life expectancy correlates better with unadjusted mortality (Figure 1A) compared to age-adjusted mortality (Figure 1B). This suggests that states with lower life expectancy experienced higher overall mortality during the pandemic, likely due to factors such as poorer healthcare infrastructure, higher prevalence of chronic conditions, and other socioeconomic differences. However, age-adjustment diminishes this effect. This finding highlights the importance of pre-pandemic health, as measured by life expectancy, in determining a state’s vulnerability to excess mortality during a pandemic. Age-adjustment partially mitigates this effect, suggesting that older populations are a significant contributing factor.

**Figure 1.**
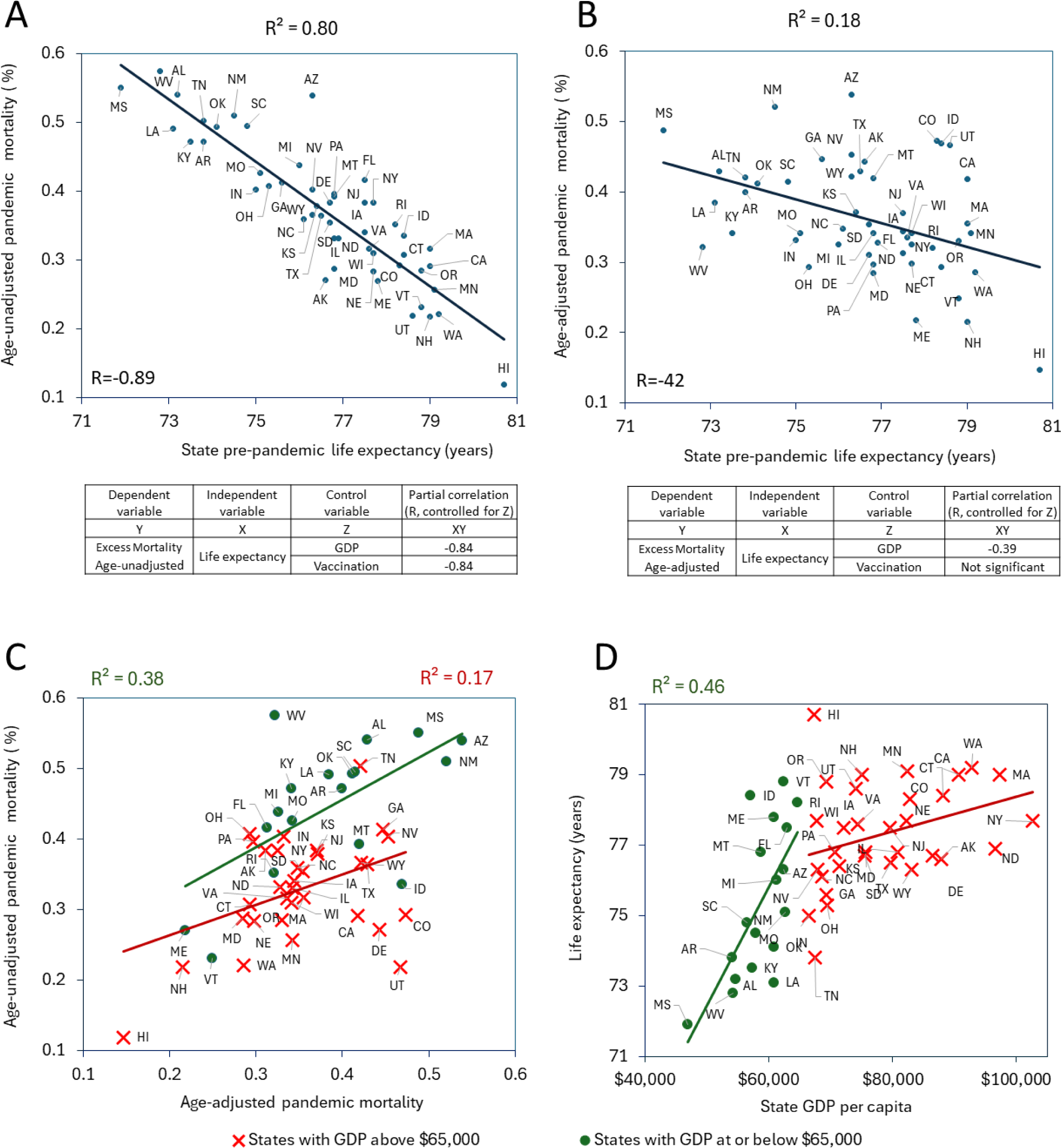
Exploring relationships between state-specific pandemic mortality, life expectancy, and GDP per capita. Mortality calculated from January 1, 2020, to July 31, 2022. The Pearson correlation coefficients displayed above the scatterplots indicate statistically significant correlations (p < 0.05). (A) Relationship of age-unadjusted pandemic mortality vs. state life expectancy. (B) Relationship of age-adjusted pandemic mortality vs. state life expectancy. (C) Relationship of age-unadjusted vs. age-adjusted state specific mortalities. (D) Relationship of state life expectancy vs GDP.

While the two mortality estimates age-adjusted and unadjusted are correlated with each other, this correlation is more pronounced for states with GDP per capita at or below $65,000 (Figure1C). This indicates that demographic factors play a substantial role in determining overall mortality in these states.

Interestingly, the strong correlation (R=-0.89, p<0.05) between pandemic age-unadjusted mortality and pre-pandemic life expectancy persists even when controlling for other variables, including vaccination and GDP, with partial correlations ranging between −0.87 and −0.84, as shown in Figure 1A. However, the partial correlation between age-adjusted mortality and life expectancy disappears when we control for vaccination rate (Figure 1B). This finding underscores the significant role of vaccination in influencing pandemic mortality outcomes, suggesting that vaccination efforts can effectively mitigate the impact of pre-existing health vulnerabilities, as reflected by life expectancy.

Additionally, not surprisingly, we noticed that states with lower GDP have lower pre-pandemic life expectancy. However, this effect is noticeable only for states with GDP at or below $65,000. Thus, the correlation between these variables is significant (p < 0.05) only for states within this subset (Figure 1D). The relationship disappears for higher income states. This suggests a diminishing effect of income on life expectancy beyond a certain GDP threshold (around $65,000 per capita in this study). Perhaps other factors besides just wealth become more important determinants of health outcomes in high-income states.

Similar to the life expectancy relationship, the link between lower pandemic mortality and higher GDP is only significant for lower-GDP states (with a GDP per capita at or below $65,000), suggesting that after reaching a certain threshold more economic resources doesn’t necessarily translate to a bigger reduction in pandemic deaths. Interestingly, there is a nuance in the relationships between GDP and pandemic mortality depending on age-consideration. Thus, within this lower-GDP subset, the correlation with GDP is weaker for age-adjusted mortality compared to unadjusted mortality (refer to Supplementary Figures 1A and 1B). This suggests that factors other than economic resources might play a more significant role in affecting mortality rates when age structure of the population is considered.

### Visualizing COVID-19 mortality waves and vaccination progress

The relationship between mortality dynamics and the COVID-19 pandemic is highlighted by the successive waves of SARS-CoV-2 infections, driven by the emergence and dominance of distinct viral variants. Fatalities arise from both direct COVID-19 impact and broader effects. The interplay between the evolving viral genomes and host responses elucidates temporal patterns in morbidity and mortality observed across pandemic infection waves. Figure 2A visually depicts daily reported COVID-19 deaths, illustrating distinct waves of infection and periods dominated by specific variants. Supplementary Figure demonstrates the chronological dominance of these variants.

**Figure 2.**
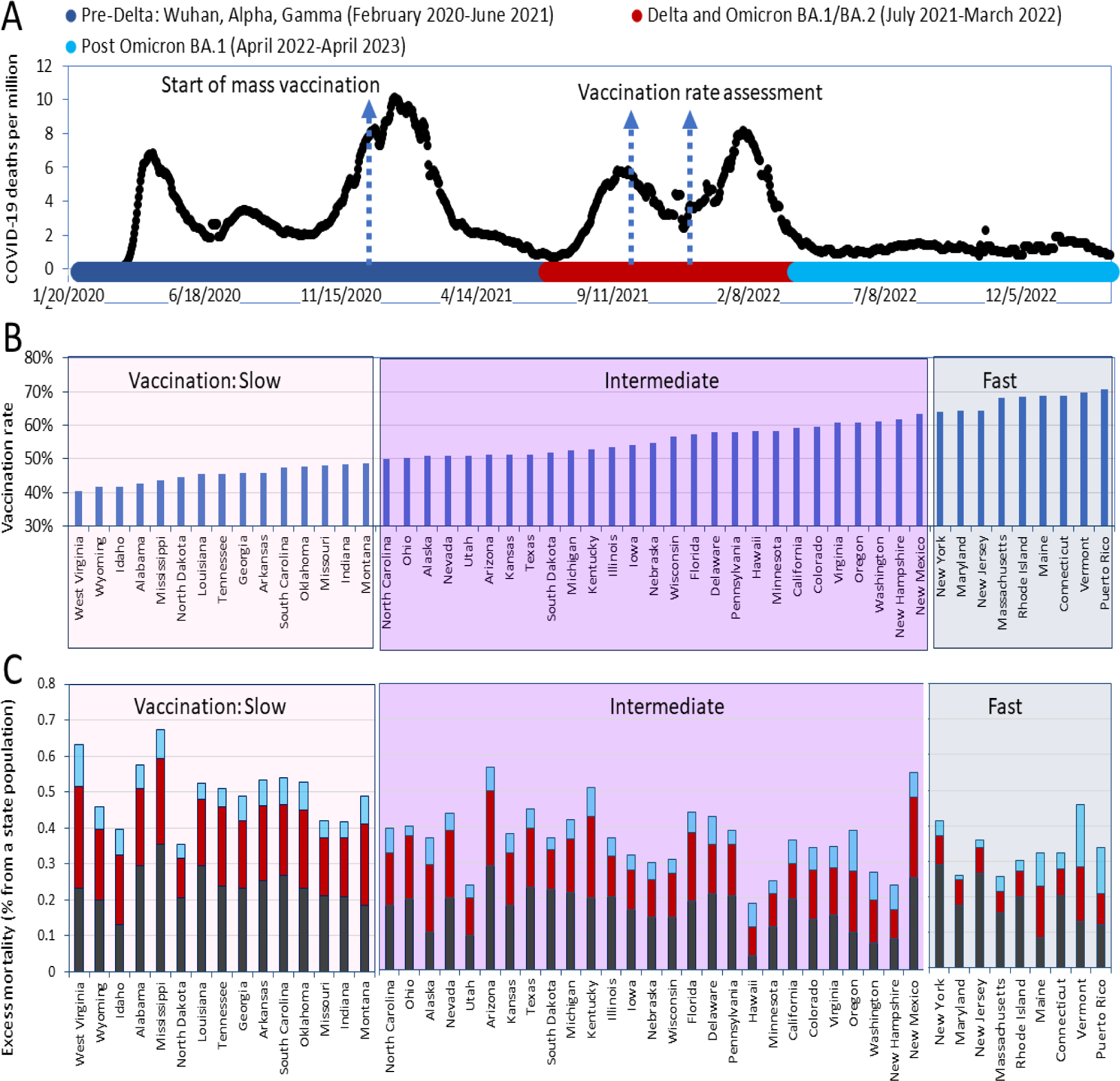
Visualization of pandemic mortality waves/periods and vaccination rates. (A) A timeline of daily diagnosed COVID-19 deaths was created using the OWID database and presented along with a scheme of the dominant virus variants causing infections. (B) States were categorized into three groups based on their vaccination coverage in ascending order by October 2021. The “fast” category (light blue background) consists of states that had achieved a vaccination rate of 60% for residents by October 2021 and 70% by January 2022. The “slow” category (light pink background) includes states in which more than half of the residents remained unvaccinated as of October 2021. The remaining states are placed in the intermediate category (with a purple background). All states and Puerto Rico are shown in the order of the increasing percentage of the state population that had received the primary series of the vaccine as of October 2, 2021. (C) Pandemic excess mortality not adjusted for age relative to the state’s population in different pandemic periods.

As a first step toward understanding pandemic disease dynamics and evaluating the effectiveness of vaccination efforts, excess mortality (non-adjusted for age) was calculated separately for the pre-Delta, Delta-Omicron BA.1, and post-Omicron BA.1 periods. These periods are marked with distinct colors in Figure 2A. In the next step of the analysis, we ranked the states according to their vaccination rates and divided them into three categories, as shown in Figure 2B. The “fast” category included states in which the vaccination coverage for residents reached 60% by October 2, 2021, and 70% by January 2, 2022. This category includes the states of Vermont, Connecticut, Maine, Rhode Island, Massachusetts, New Jersey, Maryland, and New York. The “slow” category includes states where more than half of the residents remained unvaccinated as of October 2, 2021. This category includes North Carolina, Montana, Indiana, Missouri, Oklahoma, South Carolina, Arkansas, Georgia, Louisiana, Tennessee, North Dakota, Mississippi, Alabama, Idaho, Wyoming, and West Virginia. The remaining states were categorized as having an intermediate vaccination rate.

States with higher pre-pandemic life expectancy also tended to have higher vaccination rates during the pandemic. Two parameters are highly and significantly correlated with each other (R=0.66, p<0.05). This relationship may reflect underlying factors such as better healthcare infrastructure, public health policies, and health awareness that contribute to both higher life expectancy and greater vaccine uptake.

Finally, we integrated information pertaining to mortality and vaccination rates by calculating and illustrating the number of fatalities in each state based on its respective category and time intervals (Figure 2C).

### Pronounced differences in excess mortality among state categories throughout successive pandemic periods

#### States with higher vaccination rate tended to have lower mortality rates

The interaction between excess mortality and vaccination rates throughout the pandemic were analyzed using data from different state categories across three distinct time periods. During the first pre-Delta period (time interval I lasted from January 1, 2020, to June 30, 2021), mass vaccination had just begun, and thus, did not significantly prevent excess mortality. In the second period, combining Delta and Omicron BA.1 infection waves (time interval II lasted from July1, 2021 to March 30, 2022), vaccination became widespread, potentially playing a significant role in reducing excess mortality rates. In the third period, after the Omicron BA.1 wave (time interval III lasted from April 1, 2022, to April 30, 2023), the bulk of the population acquired immunity through direct COVID-19 infection, vaccination, or hybrid immunity (a combination of both). As a result, the association between high vaccination rates and low mortality, if previously significant, may become weak or disappear altogether.

We employed odds ratio analysis to compare the likelihood of death in one category of states to another and to quantify the association between vaccination rates and mortality. The results of our analysis, conducted over distinct periods, are consistent with our predictions, and are presented in Figure 3. Initially, during the pre-Delta period, the differences between states with the slowest and fastest vaccination rates were significant, but not very pronounced (OR=1.41, 95% CI 1.32–1.5, *p*<0.01), boxplots in Figure 3A illustrate this difference for state categories created according to vaccination rates. However, as widespread mass vaccination took place during the Delta period, the disparities between states with the slowest and fastest vaccination rates became more prominent (OR=1.8, 95% CI 1.66–1.92, p<0.01), as demonstrated in Figure 3B.

**Figure 3.**
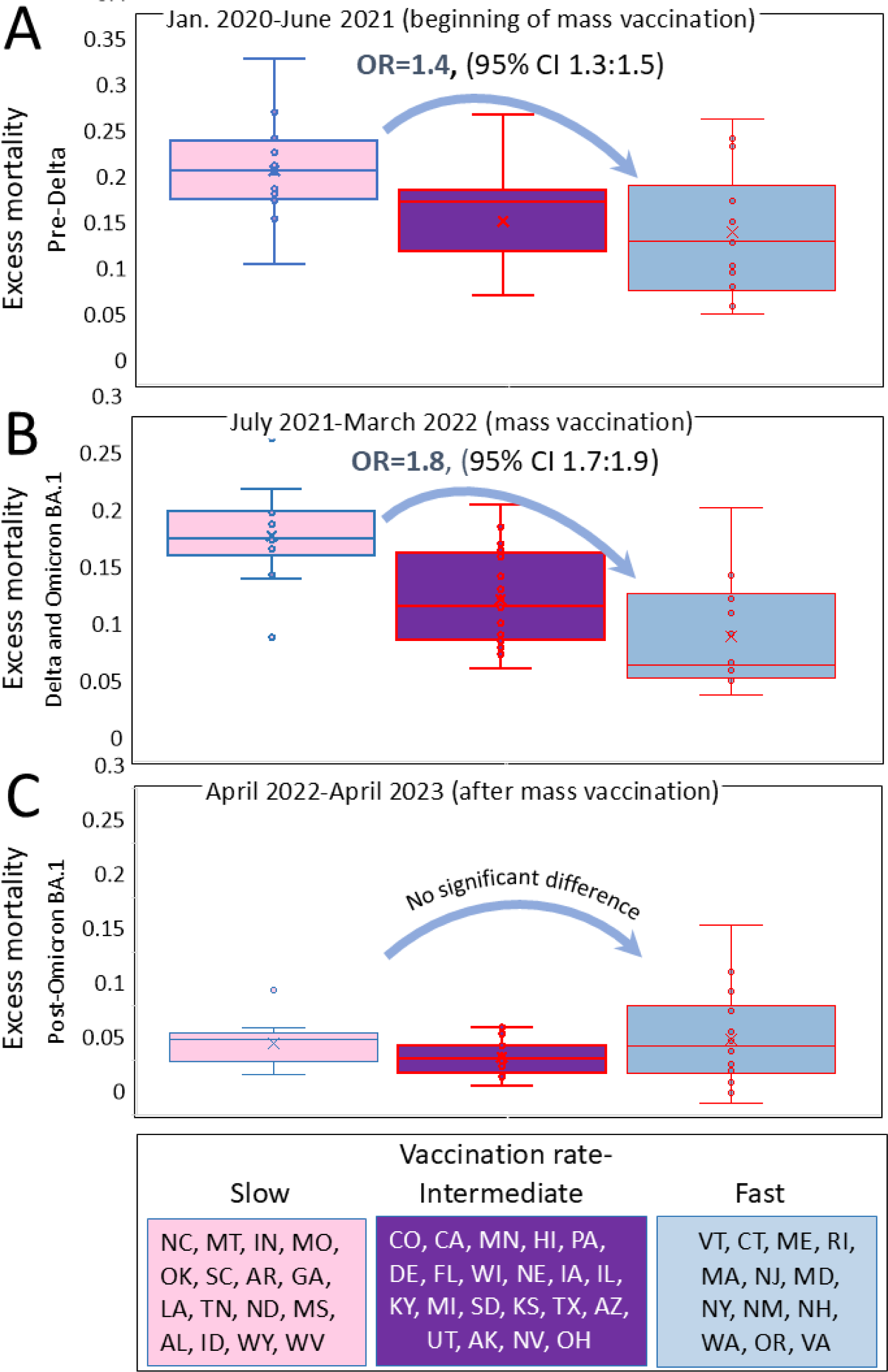
Excess mortality distributions in states categorised according to vaccination rates. States were categorized in the same manner as described in the legend of Figure 2, and each state’s excess mortality estimate (not adjuste for age) was plotted withing each category: (A) For the pre-Delta period, which corresponds to the initial phase of vaccination. (B) For the Delta and Omicron BA.1 period, representing an advanced stage of vaccination. (C) For the post-Omicron BA.1 period.

Importantly, no substantial differences in excess mortality were observed between state categories in the post-Omicron BA.1 period, when a significant proportion of the population across all states likely attained seroconversion through the combination of vaccination and prior COVID-19 infection (Figure 3C).

#### States with higher vaccination rates tend to have higher GDP per capita and longer life expectancies

In the previous subsection, we demonstrated that states with faster vaccination rates have lower excess mortality, with the most pronounced effect observed during the period of the pandemic dominated by the Delta and Omicron BA.1 variants. However, this reduction in mortality cannot be attributed solely to differences in vaccination rates. As shown in Figure 4, these states differ significantly in GDP per capita (Figure 4A) and life expectancy (Figure 4B). States with higher vaccination rates tend to have higher GDP and longer life expectancy, both of which are significantly negatively correlated with mortality. Therefore, the individual and combined contributions of these factors in determining excess mortality should be considered, weighted, and studied both together and independently.

**Figure 4.**
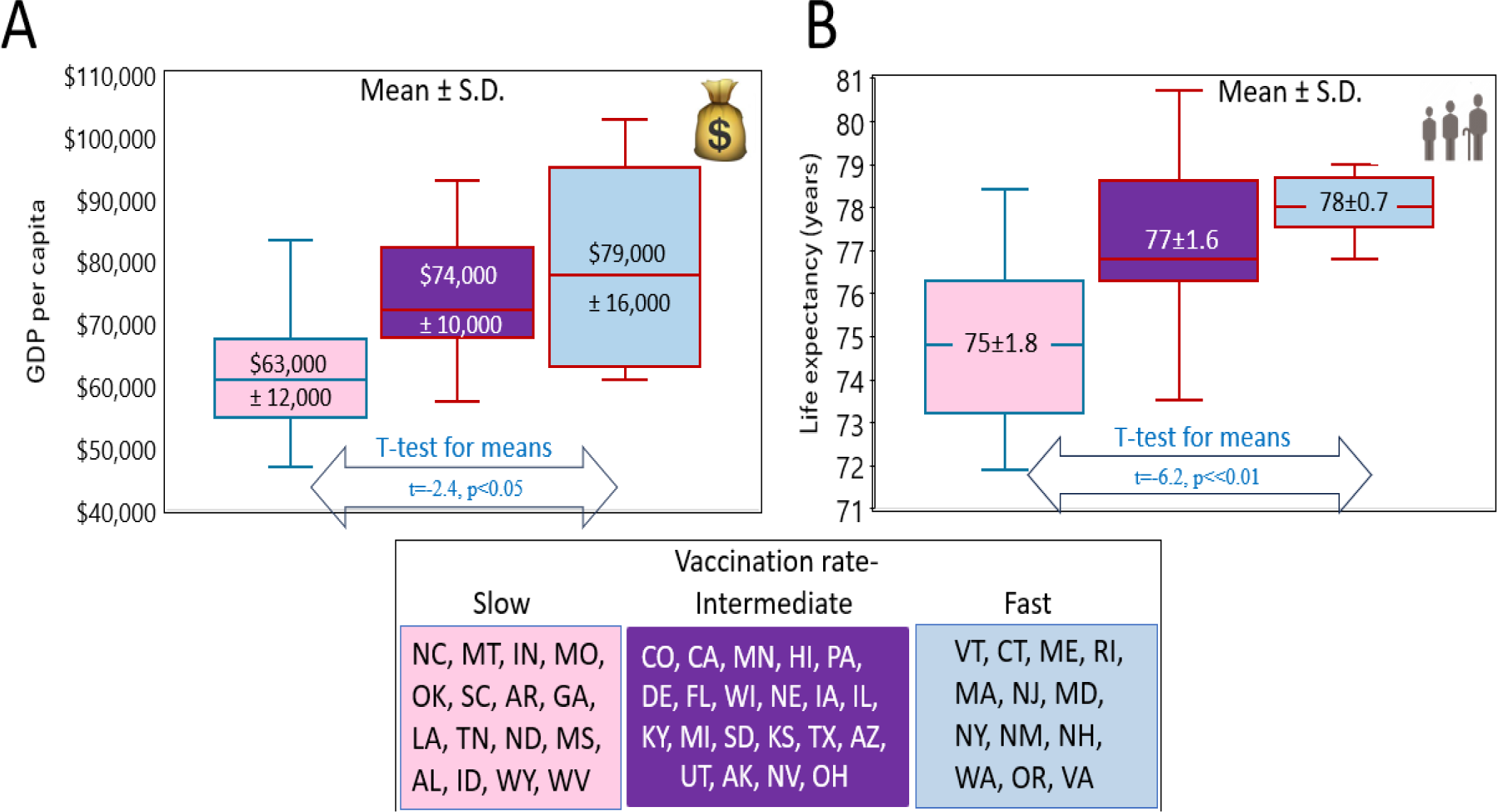
The distribution of GDPs per capita and life expectancy in states categorized as fast, intermediate, and slow according to vaccination rate. States were categorized in the same manner as described in the legend of Figure 2. GDP and life expectancy were estimated for each state within each category and resuling estimates are presented as box-plots. Means with standard deviation are shown for each category in the relevant box-plots. (A) The distribution od states according to GDP per capita, (B) The distribution according to life expectancy at birth.

#### A complex interplay between vaccination and economic status of U.S. states

Fewer excess deaths happened in states with higher GDP per capita and higher vaccination rates. Figure 5 illustrates this effect for excess mortality during the combined Delta and Omicron BA.1 variant periods, while Supplementary Figure 3 shows this effect separately for the Delta and Omicron BA.1 variants. This result is not surprising, as states with higher incomes achieved faster vaccination rates, and the relationship between these factors is significant (R=0.4, p<0.05). Both lower vaccination rates and lower GDP drive higher mortality across states; however, there are nuances in these trends.

**Figure 5.**
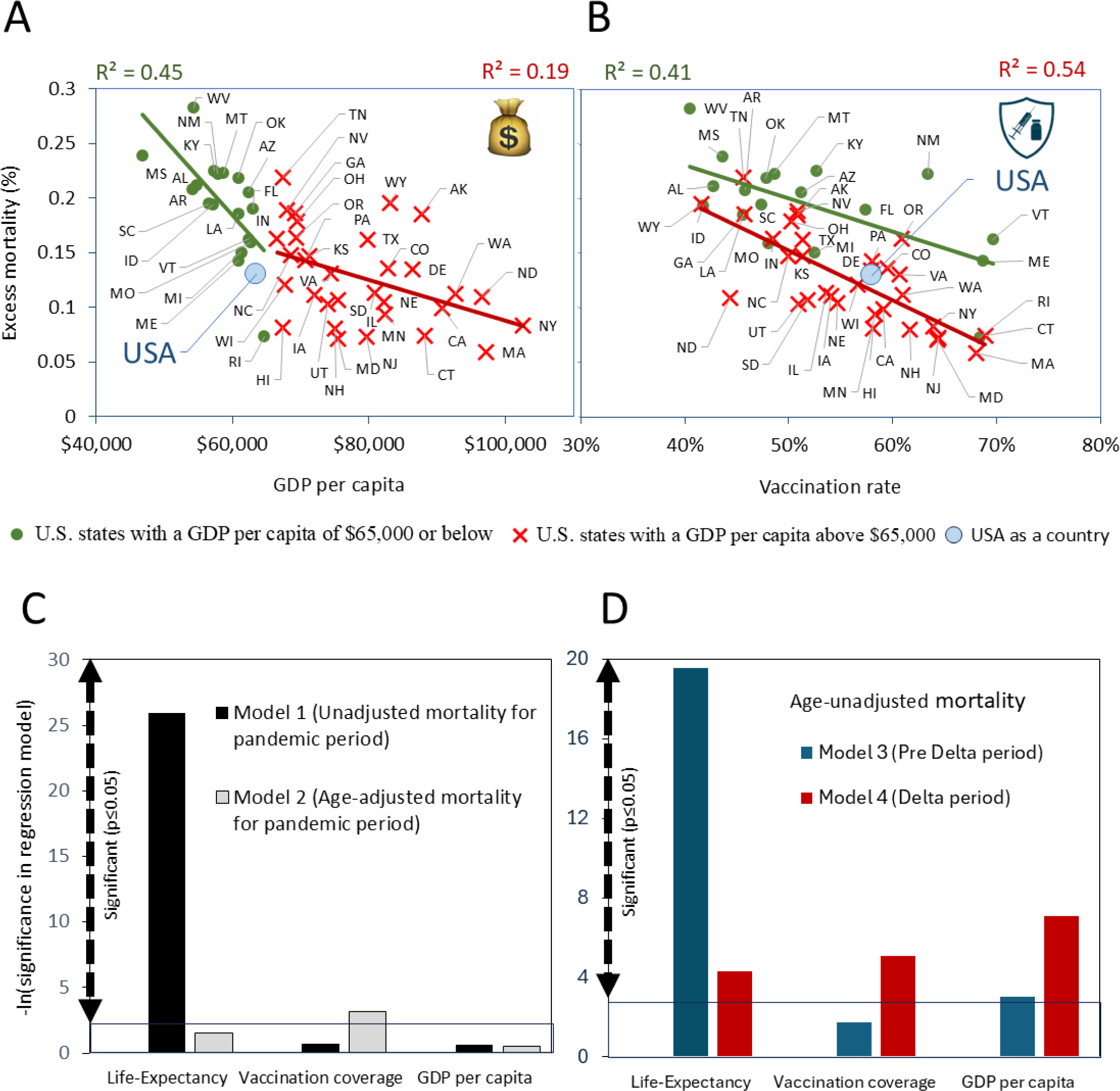
Regression analysis of pandemic excess mortality and key parameters that affected it. The top panels display results of univariable regression analysis as scatter plots. Pearson correlation coefficients, indicating significant relationships (p < 0.05), are shown above the plots. Here, the dependent variable is age-unadjusted excess mortality, with independent variables being: (A) Real GDP per capita for 2020. (B) Vaccination rate, as the percentage of the population fully vaccinated with the primary series by October 2, 2021. The bottom panels present results of multivariable regression analysis as vertical columns, reflecting negative log-transformed significance values of three key input parameters. In all models, pandemic mortalities are the dependent variables. while and life expactancy, vaccination coverage, and annual GDP per capita are independent vcariables. For these models: (C) Mortality for all pandemic periods combined, adjusted for age in model 1, and unadjusted for age in model 2. (D) Age-unadjusted mortality for two pandemic periods: Model 4 represents the pre-Delta and pre-mass vaccination period of the pandemic, while Model 5 represents the Delta variant dominance and mass vaccination period.

Among the states with GDP per capita at or below $65,000, age-unadjusted excess mortality has a stronger negative correlation with state income compared to those with higher income (Figure 5A). Moreover, with the same level of low vaccination, states with GDP above $65,000 have lower mortality compared to their less wealthy counterparts (Figure 5B). Two factors high vaccination rates and high income appear to have synergetic effects on lowing mortality across states. This synergy is also evident in multivariable regression analysis, where age unadjusted pandemic mortality is the dependent variable while vaccination rate, and GDP are independent variables. Both parameters remain significant (Supplementary Table 2).

However, age adjustment for estimates of mortality changes the picture: the synergy of two parameters in defining excess mortality disappears. The same level of low vaccination corresponds to the same level of excess deaths, regardless of GDP (compare Supplementary Figure 4 A and Figure 4 B). Thus, a high vaccination rate is the most crucial factor in defining low age-adjusted excess mortality, overriding the impact of GDP.

#### The importance and significance of mortality predictors changes troughtout the pandemic periods

Multivariable regression models indicated that life expectancy, vaccination coverage, and GDP significantly influenced age-unadjusted mortality that was estimated for all pandemic periods combined (Figure 5C). However, the relative importance of these factors shifted as the pandemic progressed. Early on, before widespread vaccination, pre-pandemic life expectancy was the strongest predictor of excess deaths (Figure 5D). With the emergence of the Delta variant and increased vaccination rates, vaccination coverage became the dominant factor, while the influence of life expectancy diminished (Figure 5D). This shift underscores the critical role of vaccination in reducing excess mortality during later stages of the pandemic, surpassing the impact of pre-existing health conditions. Notably, GDP remained a significant factor throughout the pandemic, with its importance increasing after the Delta variant’s emergence.

However, for mortality that was estimated for all pandemic periods combined, and was adjusted for age, only vaccination coverage is a factor that is a significant contributor in a multivariable model, while life expectancy and GDP were not (Figure 5C).

Overall, the data emphasize the importance of vaccination in mitigating mortality but suggest that factors such as state-specific pre-pandemic life expectancy and economic status also play a role. Furthermore, the data show that age-adjusted and unadjusted excess mortality estimates are influenced by different parameters, though high vaccination rates minimize both.

## Discussion

### Age-adjusted and unadjusted excess mortality estimates: similarities, differences, and importance in evaluating pandemic impact

When comparing Figure 1A and Figure 1B, it is evident that the correlation between age-unadjusted pandemic excess mortality and state-specific life expectancy is stronger than the correlation with age-adjusted mortality. This stronger correlation can be attributed to the influence of population age structure on both life expectancy and age-unadjusted pandemic excess mortality. The shared impact of population age structure explains the stronger correlation observed. Age-adjustment standardized the mortality rates to a common age structure, allowing for a more accurate comparison between states with different age demographics. Thus, unadjusted mortality that strongly correlates with life expectancy paints a broad picture. Age-adjusted mortality provides a more nuanced view (23), weakening the correlation but still showing a connection due to underlying factors impacting both measures.

Examining both unadjusted and age-adjusted mortality paints a more nuanced picture of the pandemic’s impact. Unadjusted mortality provides a raw count of lives lost, crucial for understanding the overall burden on each state. Age-adjusted mortality, however, helps us assess how effectively states managed the pandemic relative to their population’s vulnerabilities. As Figure 1C illustrates, these mortality estimates are correlated, likely due to factors like healthcare access, poverty, and lifestyle choices. These factors can influence both mortality rates and life expectancy. If a state has poor healthcare, we expect higher excess mortality rates and a lower life expectancy. So, even when adjusted for age structure, these underlying factors will still contribute to a connection between the two estimates. However, such underlying factors impacting both estimates are more pronounced in states with lower GDP. In contrast, states with higher GDP often have better healthcare systems and social safety nets, leading to lower mortality rates across all age groups.

It is not surprising that in the category of states with GDP below the certain threshold life expectancy correlates with state economic well-being as illustrated in Figure 1 D. For states with GDP exceeding $65,000 per capita, the correlation between economic well-being and life expectancy weakens, suggesting limited additional life expectancy gains from further economic growth.

Our current study results related to US states align with a similar pattern observed in our previous analysis of European countries (11). This pattern reveals that the relative importance of GDP and vaccination rates in predicting age unadjusted excess mortality shifted with economic context. In countries or states with lower income levels, GDP played a more prominent role, compared to its role in countries or states with higher economic status. However, age-adjusted mortality correlates and therefore likely depends only on vaccination rates. The higher vaccination rates the lower mortality estimates.

Why do countries or states with lower economic capacity demonstrate higher age-unadjusted pandemic-associated fatalities, even if they have similarly high vaccination coverage as other countries or states? A plausible explanation for this phenomenon is that states with lower GDPs, compared to those with higher GDPs, had fewer effective measures to complement vaccination in preventing COVID-19 infections. The deficiency in these measures contributed to the higher mortality rates observed in these states.

### Vaccination rates strongly correlate with pre-pandemic life expectancy across US states

Several contributing factors explain this correlation. Both metrics reflect a state’s healthcare system capacity and its ability to adapt to crises. Additionally, strong public trust and a population willing to comply with public health measures play a role. Indeed, pre-pandemic life expectancy is highly and significantly inversely correlated with vaccination hesitancy estimated during the pandemic. This suggests a form of synergy between state governments and their residents. Effective government action to combat the pandemic is met with greater public cooperation, including low vaccine hesitancy that resulted in high vaccination uptake.

### Association between high vaccination coverage and excess mortality disappeared following the dominance of Omicron BA.1 virus variant

Our analysis demonstrates a progressively stronger association between low vaccination rates and high excess mortality that peaked during the Delta period. Eventually, death rates converge across states in the post-Omicron BA.1 period, due to widespread seroconversion from natural infections. In the period following the dominance of Omicron BA.1, a considerable proportion of the population in all states had already achieved immunity, which in states with low vaccination rates was offset by high rates of natural infections. These infections resulted in excess deaths, but also contributed to a strengthening of population immunity and this strengthening may have been substantial because of the considerable number of infections that had occurred by the end of March 2022 in most states. Highly likely that all states had achieved a comparable level of immunity by that point.

The findings from our analysis of both European countries and U.S. states offer insights for shaping strategies in future pandemics or infections surges of a lesser degree. Understanding the patterns observed can inform proactive measures to mitigate the impact, particularly in regions with lower economic capacities.

## Conclusions

- States with lower life expectancy tend to have much higher pandemic-related deaths.
- **Economic well-being (GDP) influences life expectancy, but with a potential threshold effect**. In this study the GDP threshold was defined as a value of $65,000 per capita for 2020. Once a state achieves a certain level of economic well-being, its life expectancy does not change significantly with further increases in GDP.
- **Economic well-being (GDP) also plays a role in reducing excess mortality that is not adjusted for age**, but its importance varies depending on state income level. In states with GDP per capita below the threshold, there’s a stronger negative correlation between GDP and mortality, while in remaining states, vaccination rates have a more prominent effect.
- **Vaccination rates play a most significant role in reducing excess mortality.** This effect is consistent across states regardless of income level. Notably, in multivariable regression models, only vaccination rate, and not GDP remained as a significant factor affecting age-adjusted pandemic mortality. The category of front-runner states with the fastest vaccination campaigns that other states can model includes Vermont, Connecticut, Maine, Rhode Island, Massachusetts, New Jersey, Maryland, and New York.

## Data Availability

Our work was performed with publicly available datasets. All data generated or analyzed during this study is provided as supplementary information files.

## Author Contributions

OM - data curation, formal analysis, research, writing, figures design; AO - statistical analysis, conceptualization; SS - conceptualization, statistical analysis, methodology, writing. All authors contributed to the article and approved the submitted version.

---

**Supplementary Figure 1.**
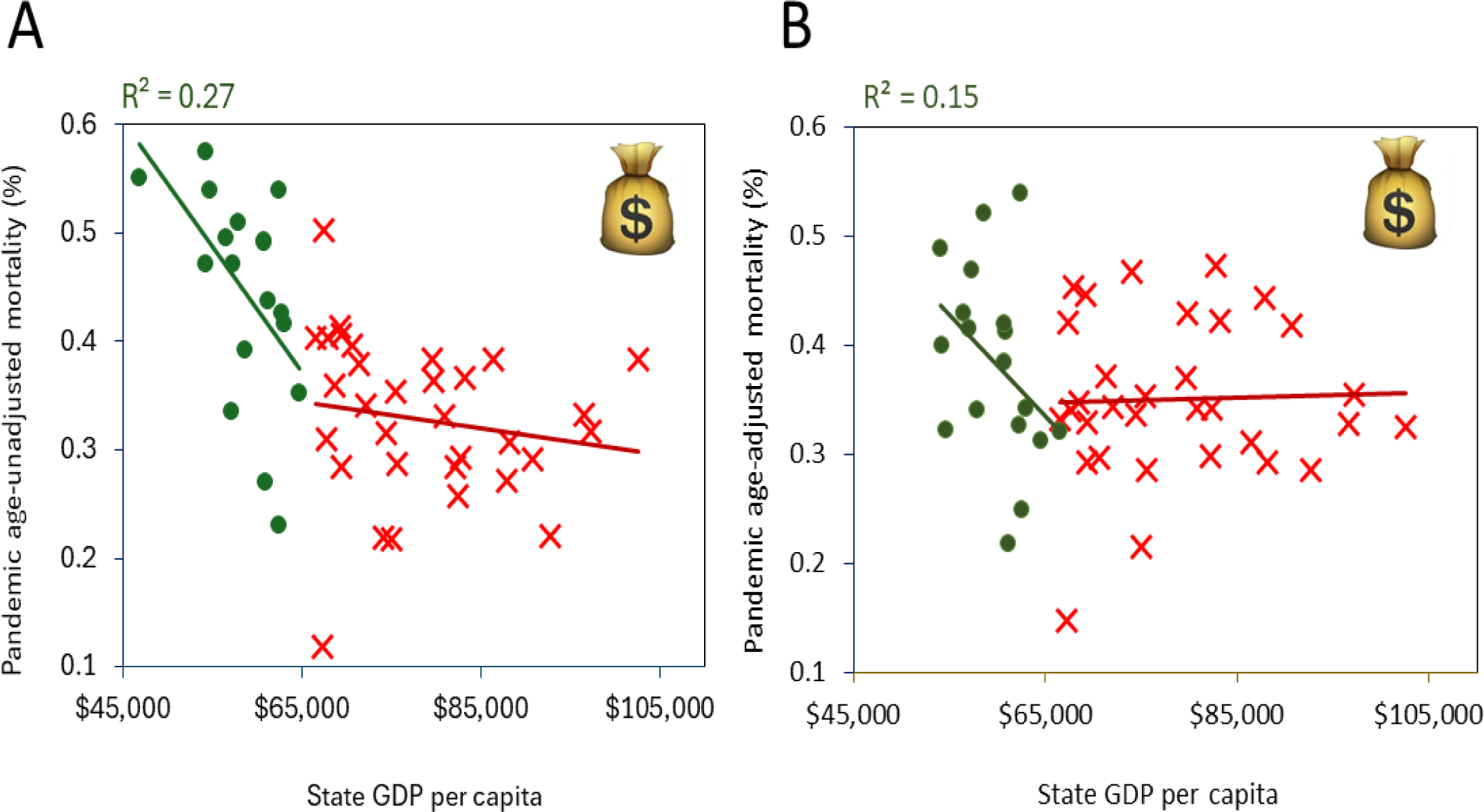
Relationships between state-specific pandemic mortality, and GDP per capita. Excess mortality was calculated as the percentage of normalized excess deaths for the state population from January 1, 2020, to July 31, 2022. The Pearson correlation coefficients displayed above the scatterplots indicate statistically significant correlations (p < 0.05). (A) Relationship of age-unadjusted pandemic mortality vs. GDP (B) Relationship of age-adjusted pandemic mortality vs. GDP. of age-unadjusted vs. age-adjusted state specific mortalities.

**Supplementary Figure 2.**
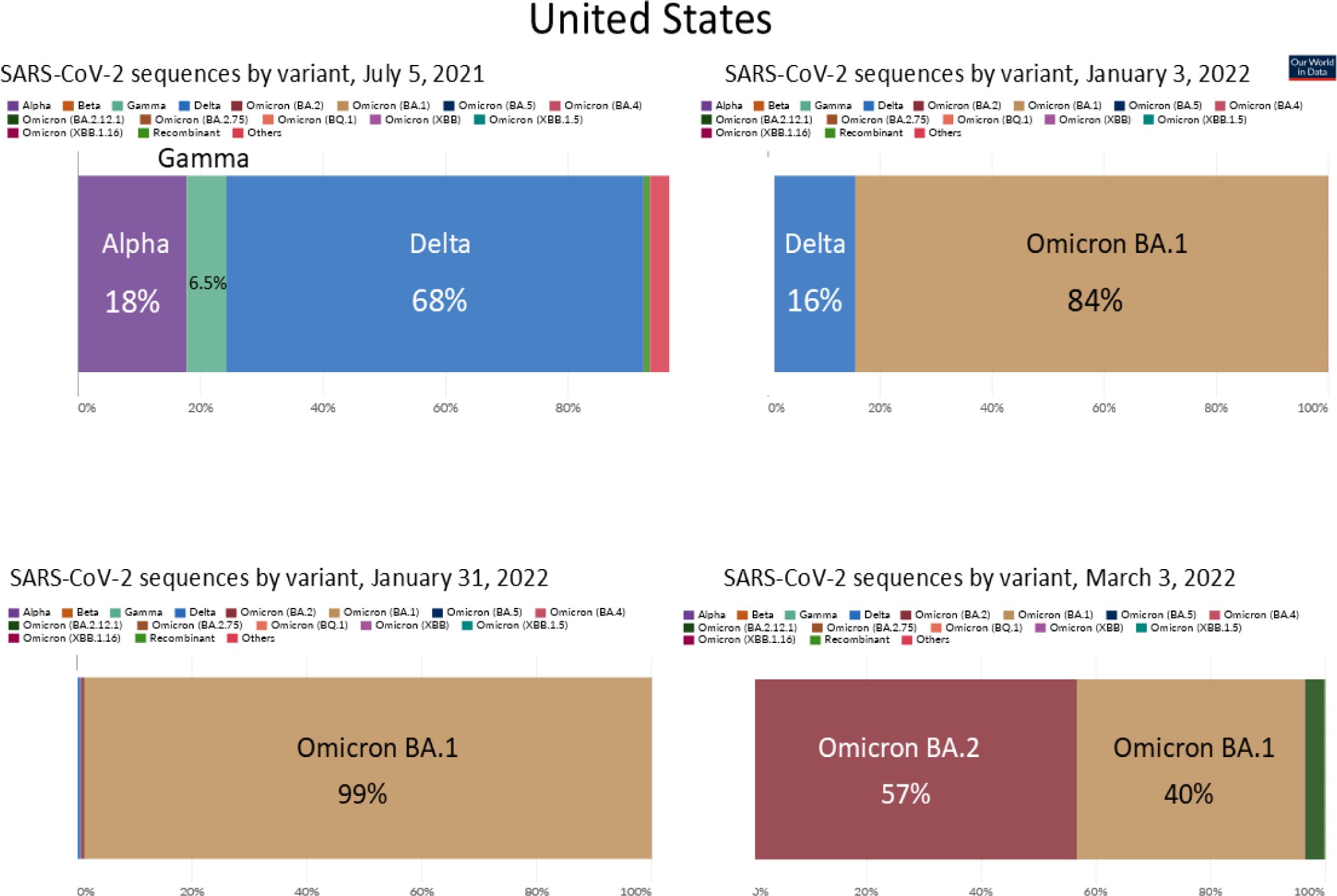
Chronological Emergence of SARS-CoV-2 Dominant Variants in the USA.

**Supplementary Figure 3.**
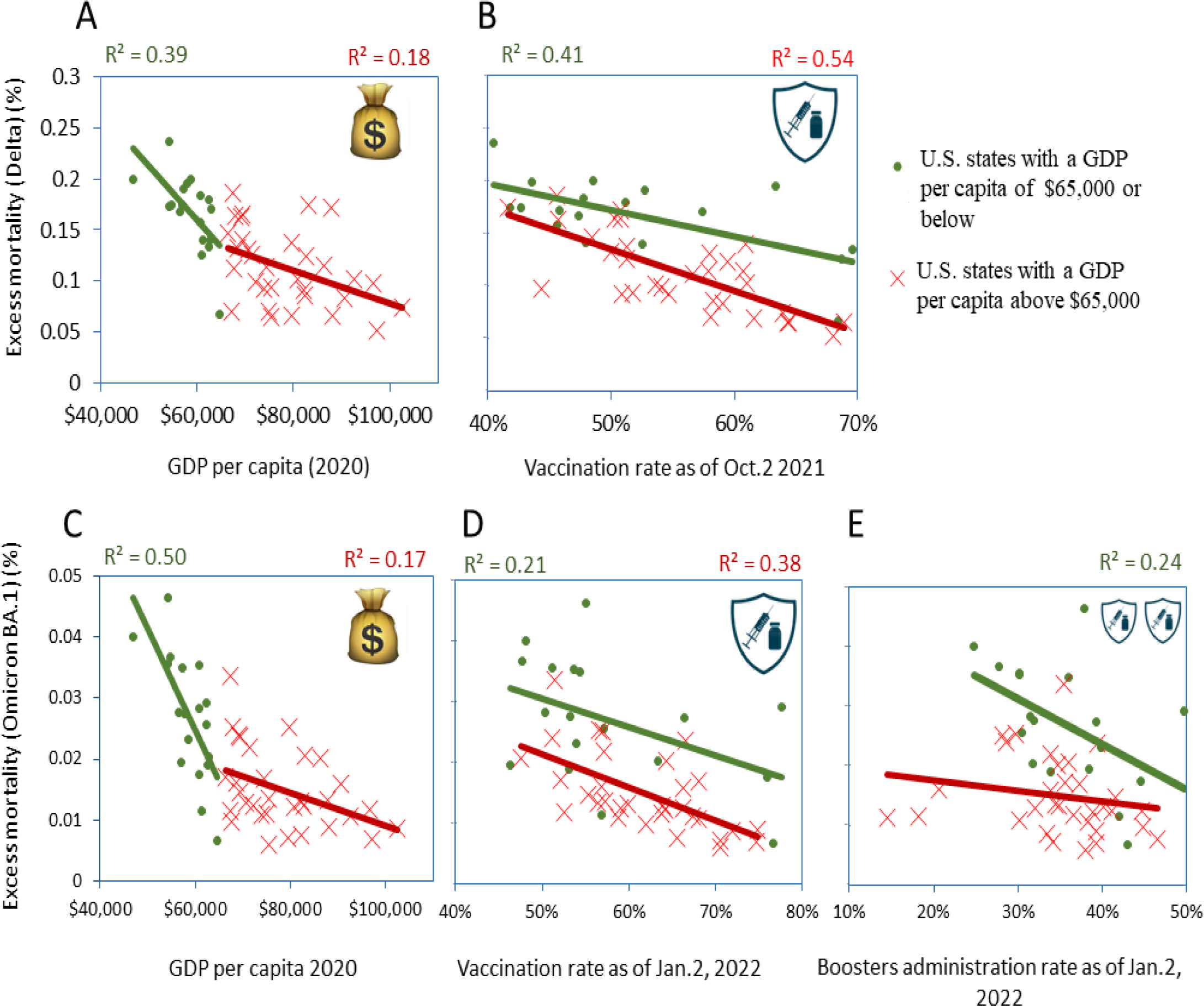
Correlation analysis of pandemic excess mortality for Delta and Omicron BA.1 periods separatly. Only statistically significant correlations (p < 0.05) are presented above the corresponding scatter plots. Excess mortality that was not adjusted for age is depicted for: (A) Delta period in relation to the annual actual GDP per capita in 2020. (B) Delta period in relation to vaccination rates, expressed as a percentage of the state’s fully vaccinated population as of October 2, 2021. (C) Omicron BA.1 period in relation to GDP per capita. (D) Omicron BA.1 period in relation to vaccination rates, calculated as a percentage of the state’s fully vaccinated population as of January 2, 2022. (E) Omicron BA.1 period in relation to booster administration rates, calculated as a percentage of the state’s population that received an additional dose as part of the primary vaccine series as of January 2, 2022.

**Supplementary Figure 4.**
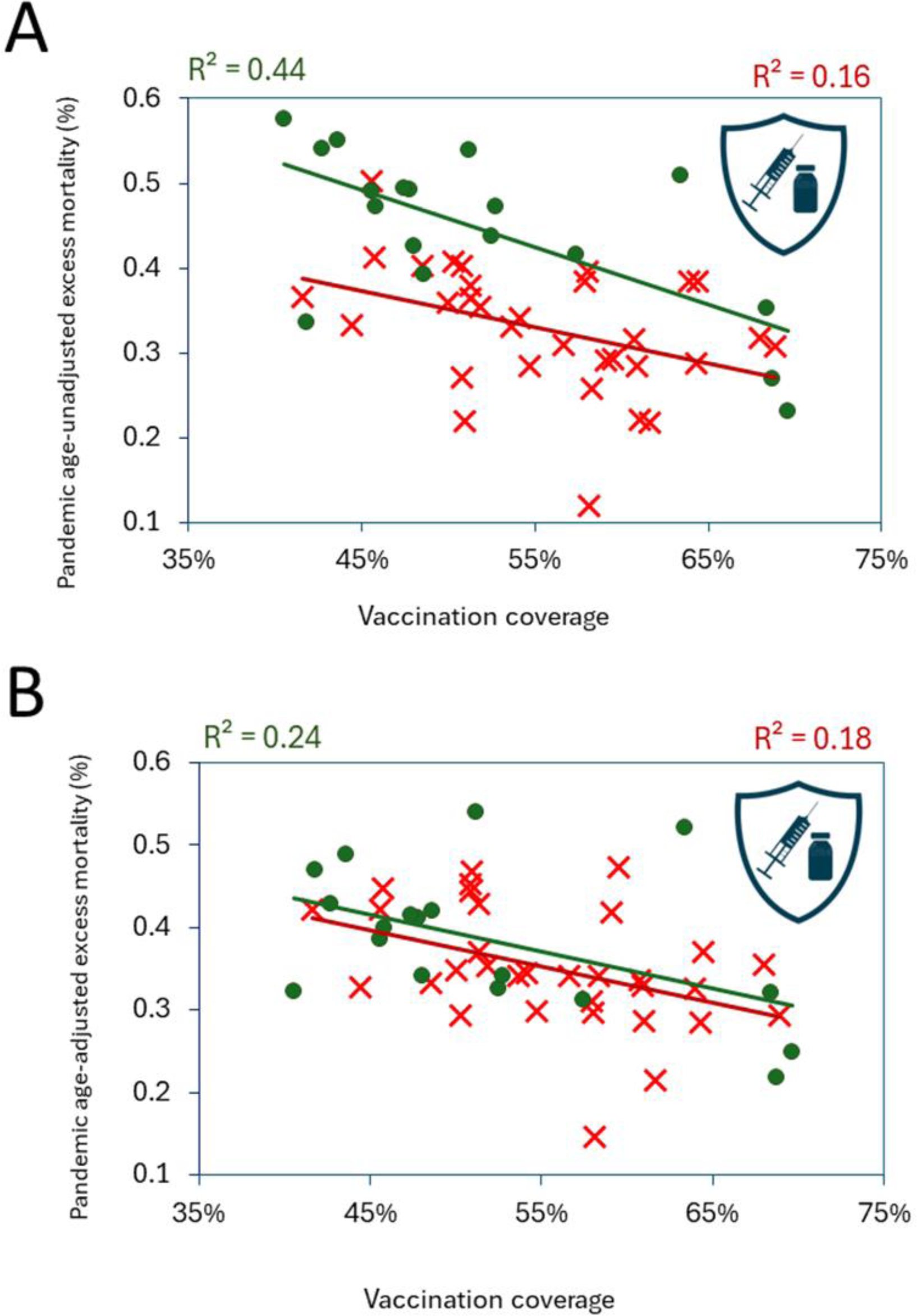
Comparison of relationships of vaccination coverages with pandemic excess mortality estimated with and without adjustement for age. Both estimates were perforemd for the period spanning from January 1, 2020, to July 31, 2023. State vaccination coverage was estimated as October 2, 2021. Pearson correlation coefficients reflecting significant relationships (p < 0.05) are shown above the scatter plots. (A) The dependent variable is excess mortality,not adjusetd for age. (B) The dependent variable is age-adjusted excess mortality.

